# A systems approach to inflammation identifies therapeutic targets in SARS-CoV-2 infection

**DOI:** 10.1101/2020.05.23.20110916

**Authors:** Frank L. van de Veerdonk, Nico A.F. Janssen, Inge Grondman, Aline H. de Nooijer, Valerie A.C.M. Koeken, Vasiliki Matzaraki, Collins K. Boahen, Vinod Kumar, Matthijs Kox, Hans J.P.M. Koenen, Ruben L. Smeets, Irma Joosten, Roger J.M. Brüggemann, Ilse J.E. Kouijzer, Hans G. van der Hoeven, Jeroen A. Schouten, Tim Frenzel, Monique Reijers, Wouter Hoefsloot, Anton S.M. Dofferhoff, Angèle P.M. Kerckhoffs, Marc J.T. Blaauw, Karin Veerman, Coen Maas, Arjan H. Schoneveld, Imo E. Hoefer, Lennie P.G. Derde, Loek Willems, Erik Toonen, Marcel van Deuren, Emeritus Jos W.M. van der Meer, Reinout van Crevel, Evangelos J. Giamarellos-Bourboulis, Leo A.B. Joosten, Michel M. van den Heuvel, Jacobien Hoogerwerf, Quirijn de Mast, Peter Pickkers, Mihai G. Netea, on behalf of the RCI-COVID-19 study group

**Affiliations:** Department of Internal Medicine and Radboud Center for Infectious Diseases, Radboud University Medical Center, 6500 HB Nijmegen, The Netherlands; Department of Genetics, University Medical Center Groningen, 9700 CC Groningen, the Netherlands; Department of Intensive Care Medicine and Radboud Center for Infectious Diseases, Radboud University Medical Center, 6500 HB Nijmegen, the Netherlands; Laboratory Medicine, Laboratory for Medical Immunology, Radboud University Medical Center, 6500 HB Nijmegen, the Netherlands; Radboudumc Laboratory for Diagnostics, Radboud University Medical Center, 6500 HB Nijmegen, the Netherlands; Department of Pharmacy, Radboud University Medical Center, 6500 HB Nijmegen, The Netherlands; Department of Pulmonary Diseases, Radboud University Medical Center, 6500 HB Nijmegen, The Netherlands; Department of Internal Medicine, Canisius Wilhelmina Hospital, 6500 GS Nijmegen, the Netherlands; Department of Nephrology, Jeroen Bosch Ziekenhuis, ’s-Hertogenbosch, the Netherlands; Department of Geriatric Medicine, Jeroen Bosch Hospital, 5200 ME ’s-Hertogenbosch, the Netherlands; Department of Internal Medicine, Bernhoven Hospital, 5400 AS Uden, The Netherlands; Department of Internal Medicine, Sint Maartenskliniek, 6500 GM Nijmegen, the Netherlands; Central Diagnostic Laboratory, University Medical Center Utrecht, 3508 GA Utrecht, the Netherlands; Department of Intensive Care Medicine, University Medical Center Utrecht, 3508 GA Utrecht, The Netherlands; R&D Department, Hycult Biotechnology, 5405 PB Uden, the Netherlands; 4^th^ Department of Internal Medicine, National and Kapodistrian University of Athens, 124 62 Athens, Greece; Immunology and Metabolism, Life & Medical Sciences Institute, University of Bonn, 53115 Bonn, Germany

**Keywords:** COVID-19, Inflammation, Pathogenesis, Cytokines, Complement, Bradykinin, ARDS, Proteomics

## Abstract

**Background:** Infection with SARS-CoV-2 manifests itself as a mild respiratory tract infection in the majority of individuals, which progresses to a severe pneumonia and acute respiratory distress syndrome (ARDS) in 10-15% of patients. Inflammation plays a crucial role in the pathogenesis of ARDS, with immune dysregulation in severe COVID-19 leading to a hyperinflammatory response. A comprehensive understanding of the inflammatory process in COVID-19 is lacking.

**Methods:** In this prospective, multicenter observational study, patients with PCR-proven or clinically presumed COVID-19 admitted to the intensive care unit (ICU) or clinical wards were included. Demographic and clinical data were obtained and plasma was serially collected. Concentrations of IL-6, TNF-α, complement components C3a, C3c and the terminal complement complex (TCC) were determined in plasma by ELISA. Additionally, 269 circulating biomarkers were assessed using targeted proteomics. Results were compared between ICU and non ICU patients.

**Findings:** A total of 119 (38 ICU and 91 non ICU) patients were included. IL-6 plasma concentrations were elevated in COVID-19 (ICU vs. non ICU, median 174.5 pg/ml [IQR 94.5-376.3] vs. 40.0 pg/ml [16.5-81.0]), whereas TNF-α concentrations were relatively low and not different between ICU and non ICU patients (median 24.0 pg/ml [IQR 16.5-33.5] and 21.5 pg/ml [IQR 16.0-33.5], respectively). C3a and terminal complement complex (TCC) concentrations were significantly higher in ICU vs. non ICU patients (median 556.0 ng/ml [IQR 333.3-712.5]) vs. 266.5 ng/ml [IQR 191.5-384.0] for C3a and 4506 mAU/ml [IQR 3661-6595] vs. 3582 mAU/ml [IQR 2947-4300] for TCC) on the first day of blood sampling. Targeted proteomics demonstrated that IL-6 (logFC 2.2), several chemokines and hepatocyte growth factor (logFC 1.4) were significantly upregulated in ICU vs. non ICU patients. In contrast, stem cell factor was significantly downregulated (logFC −1.3) in ICU vs. non ICU patients, as were DPP4 (logFC −0.4) and protein C inhibitor (log FC −1.0), the latter two factors also being involved in the regulation of the kinin-kallikrein pathway. Unsupervised clustering pointed towards a homogeneous pathogenetic mechanism in the majority of patients infected with SARS-CoV-2, with patient clustering mainly based on disease severity.

**Interpretation:** We identified important pathways involved in dysregulation of inflammation in patients with severe COVID-19, including the IL-6, complement system and kinin-kallikrein pathways. Our findings may aid the development of new approaches to host-directed therapy.

**Funding:** Vidi grant (F.L.v.d.V.) and Spinoza grant (M.G.N.) from the Netherlands Organization for Scientific Research, and ERC Advanced Grant (#833247 to M.G.N.).

## Introduction

Severe acute respiratory syndrome coronavirus-2 (SARS-CoV-2) is a highly contagious virus that spread rapidly from China to the rest of a highly-interconnected world to become a pandemic in March 2020.^1^ The clinical spectrum of SARS-CoV-2 infection (also termed COVID-19) varies from asymptomatic disease and symptoms of mild upper respiratory tract infection, to severe pneumonia with acute respiratory distress syndrome (ARDS), respiratory failure and death.^2^ The spread of SARS-CoV-2 around the world infected millions of people in several months and killed tens of thousands. Effective treatments are therefore urgently needed for the high numbers of severely ill patients. Although much has been learned in a very short time, a comprehensive understanding of the pathophysiology of COVID-19 is still lacking.

The most important complication in COVID-19 is respiratory failure, which is mediated by local inflammation and edema, the development of ARDS, and subsequently hypoxia. Inflammation plays a central role in the pathogenesis of ARDS and circulating concentrations of proinflammatory cytokines such as interleukin (IL)-6, tumour necrosis factor (TNF)-α, monocyte chemoattractant protein (MCP)-1, macrophage inflammatory protein (MIP)-1α and interferon-γ inducible protein (IP)-10 are higher in COVID-19 patients on the intensive care unit (ICU) than in those who do not require ICU admission.^2^ This systemic inflammatory response is also associated with elevated D-dimer concentrations in the circulation and hyperactive CCR6+Th17+ T-cells locally in the lung.^3,4^ A recent study showed that hyperinflammation in COVID-19 patients is characterised by a high cytokine production capacity of circulating monocytes despite the severity of the disease, a feature different from other types of sepsis.^5^ The systemic inflammatory response in COVID-19 patients is accompanied by lymphopenia, which is one of the most striking features encountered in severely ill patients, with both CD4 and CD8 lymphocytes being deficient.^6^

Whereas from these data an exuberant innate immune response appears to represent the main immune dysregulation in patients with severe COVID-19 infection, so far only a limited number of inflammatory mediators known to be involved in other diseases have been assessed. A comprehensive, unbiased understanding of the inflammatory processes in COVID-19 is lacking, while this is crucial for the development of effective host-directed therapies to restore the immune balance in COVID-19 patients. In addition, it is not known whether the pathophysiology of COVID-19 is homogeneous between patients, or whether immune endotypes are present which may lead to complications through different pathophysiological mechanisms, as have been identified in bacterial sepsis patients.^7^ In the present study, we used targeted proteomics and systems biology analyses in a systems-based approach to analyze the inflammatory response in patients with mild versus severe COVID-19. We utilised a combination of multiple ELISA measurements and Olink panels to measure more than 200 different circulating inflammatory parameters in the plasma of COVID-19 patients. We subsequently identified several major inflammatory pathways that discriminate between severely ill patients and patients with mild disease, which therefore represent potential starting points for therapeutic targeting. Subsequently, the unbiased analysis of the proteomics data also suggests a homogeneous inflammatory pathogenesis of the disease, with the main stratification of patients based on disease severity, rather than different inflammatory endotypes.

## Methods

### Patient inclusion and plasma collection

This study was performed according the latest version of the declaration of Helskini and guidelines for good clinical practice. The local independent ethical committee approved the study protocol (CMO 2020-6344 and CMO 2016-2923). All patients (or their representatives) admitted to the Radboud University Medical Center (Radboudumc), a tertiary care university medical care facility, with a PCR-proven SARS-CoV-2 infection or presumed infection (based on signs and symptoms and findings on computed tomography (CT) scans) were asked for informed consent for participation in this study. After obtaining verbal informed consent, ethylenediaminetetraacetic acid (EDTA) blood was collected three times per week (ICU) or every 48 hours (non ICU wards) during times of routine venapuncture for laboratory testing and stored at 4 °C until further processing in the laboratory. After centrifugation for 10 minutes at 3800 rpm (2954 g) at room temperature, plasma was collected and stored at either −20 °C for later enzyme-linked immunosorbent assay (ELISA) for cytokines and chemokines or stored at −80 °C for later analysis. Demographic data, medical history and clinical laboratory measurements were collected from the medical file, wehere available, and processed in encoded form in electronic case report forms using Castor electronic data capture (Castor EDC, Amsterdam, the Netherlands).

For complement data analysis, data from healthy controls (from the 200FG cohort; www.humanfunctionalgenomics.org) and bacterial septic shock patients early in their course of disease (classified according to the Sepsis 3 criteria) (from the PROVIDE Study cohort; ClinicalTrials.gov NCT03332225) were used as comparisons for COVID-19 patients.

### Cytokine and chemokine ELISAs

Commercially available ELISA kits (Quantikine ELISA kits, R&D Systems, Inc., Minneapolis, MN, USA) were used for assessing concentrations of IL-6 and TNF-α in patient plasma according to the manufacturer’s instructions. Concentrations of complement system components C3a, C3c and the terminal complement complex (TCC) in patient plasma were performed by commercially available ELISA kits (Hycult Biotech, Uden, the Netherlands) according to the manufacturer’s instructions. Inter-assay variation was assessed by calculating the coefficient of variation (%CV) for the quality control samples between assay runs. A %CV of ≤ 15 was considered low variation.

## Proteomics analysis

Circulating proteins were measured in plasma using the commercially available multiplex proximity extension assay (PEA) from Olink Proteomics AB (Uppsala Sweden).^8^ In this assay, proteins are recognised by pairs of oligonucleotide-labeled antibodies (“probes”),. When the two probes are in close proximity, a new PCR target sequence is formed by a proximity-dependent DNA polymeration reaction. The resulting sequence is subsequently detected and quantified using a standard real-time PCR. In total, proteins from three different panels were measured (Olink® Inflammation, Olink® Cardiometabolic and Olink® Cardiovascular II), which resulted in the measurement of 269 different biomarkers. Proteins are expressed on a log2-scale as normalised protein expression (NPX) values, and normalised using bridging samples to correct for batch variation.

For the proteomic analyses, biomarkers were excluded from the analysis when the target protein was detected in less than 80% of the samples. Protein concentrations under the detection threshold were replaced with the proteins lower limit of detection (LOD). In addition, Olink proteomics performed quality control per sample during which samples that deviate less than 0.3 NPX from the median pass the quality control.

### Statistical analysis

For demographic, laboratory, cytokine/chemokine and complement data, ICU and non ICU groups were compared using the Mann-Whitney test or Kruskal-Wallis test with Dunn’s multiple comparison test (when comparing more than two groups), assuming non-Gaussian distribution of variables. Percentages were compared using Fisher’s exact test. A *p*-value < 0.05 was considered statistically significant. Statistical analyses were performed using either GraphPad Prism 5 for Windows (version 5.03, GraphPad Software, Inc., San Diego, CA, USA) or R/Bioconductor (https://www.R-project.org/). Differential expression (DE) analysis of Olink® proteins between ICU and non-ICU groups was performed using the R package limma,^9^ where a linear model was applied with age and sex as covariates. limma uses an empirical Bayes method to moderate the standard errors of the estimated log-fold changes. Unsupervised hierarchical clustering was performed to identify patient endotypes.

## Results

### Baseline characteristics and laboratory values of patients with COVID-19

Plasma was collected from 119 patients with confirmed or presumed (based on signs and symptoms, imaging results and epidemiological exposure) COVID-19 admitted to ICU departments (n = 38) or designated clinical wards (n = 81). Age, sex, body mass index (BMI), day of admission at the time of first blood collection and percentages of polymerase chain reaction (PCR)-proven versus presumed COVID-19 diagnosis are shown in Table 1. More men than women were admitted, and the mean BMI was 27.6 kg/m^2^ (standard deviation (SD): 4.3). Routine clinical laboratory results (available for 79/119 patients included) demonstrated that COVID-19 patients had lymphopenia with a median of 0.7 × 10^9^/l (interquartile range [IQR] 0.4-1.1). Neutrophils were higher in ICU patients (median 7.3 × 10^9^/l [IQR 4.1-9.3]) vs. 3.6 × 10^9^/l [IQR 3.0-5.3] in non ICU patients, *p* = 0.0024) and median thrombocyte counts were normal and not significantly different between ICU and non ICU patients (228 × 10^9^/l [IQR 154.3-278] vs. 185.5 × 10^9^/l [IQR 122.8-278], respectively, *p* = 0.3773). D-dimer and CRP concentrations were higher in patients admitted to the ICU compared to non ICU patients (3420 ng/ml [IQR 1890-6805] vs. 1150 ng/ml [IQR 760-1750], *p* < 0.0001 and 266.5 mg/l [IQR 149.8-308.5] vs. 79 mg/l [IQR 43-139.5], *p* < 0.0001, respectively; Table 2). Although circulating ferritin concentrations were also increased in ICU patients as compared to non ICU COVID-19 patients, no statistically significant differences were observed (1470 μg/l [IQR 747.8-1965] vs. 991 μg/l [IQR 566.5-1542], p = 0.0557; Table 2).

**Table 1.** Demographic and COVID-19 admission data for total population of patients for which cytokine/chemokine ELISAs were performed

|  | **Total (n = 119)** | **Non ICU (n = 81)** | **ICU (n = 38)** | **p (Non ICU vs. ICU)** |
| --- | --- | --- | --- | --- |
| Age (mean +/- SD) | 64.4 (+/- 12.7) | 64.5 (+/- 13.4) | 64.0 (+/- 11.3) | 0.6730 |
| Sex (n, %) |  |  |  | 0.6709 |
| Male | 84 (70.6) | 56 (69.1) | 28 (73.7) |  |
| Female | 35 (29.4) | 25 (30.9) | 10 (26.3) |  |
| BMI (kg/m<sup>2</sup>; mean +/- SD) | 27.6 (+/- 4.3)<sup>a</sup> | 27.3 (+/- 4.5)<sup>b</sup> | 28.0 (3.8) | 0.2499 |
| Days of admission at first blood sample (days; median and range) | 2.0 (0 - 15) | 2.0 (0 - 15) | 4.0 (1 - 13) | **0.0001** |
| Proven (vs. presumed) COVID-19 (n, %) | 114 (95.8) | 77 (95.1) | 37 (97.4) | 1.000 |

<sup>a</sup>n = 115; <sup>b</sup>: n = 77

**Table 2.** Population with at least assessment of a WBC at the first time point for plasma collection, values at first time point

|  | **Total (n = 79)** | **Non ICU (n = 53)** | **ICU (n = 26)** | **p (Non ICU vs. ICU)** |
| --- | --- | --- | --- | --- |
| Age (mean +/- SD) | 63.7 (+/- 13.7) | 62.6 (+/- 14.3) | 66.0 (+/- 12.4) | 0.2661 |
| Sex (n, %) |  |  |  | 0.2150 |
| Male | 53 (67.1) | 33 (62.3) | 20 (76.9) |  |
| Female | 26 (32.9) | 20 (37.7) | 6 (23.1) |  |
| BMI (kg/m<sup>2</sup>; mean +/- SD) | 27.3 (+/- 4.4)<sup>a</sup> | 27.1 (4.6)<sup>b</sup> | 27.7 (4.1) | 0.4235 |
| Days of admission at first blood (days; median and range) | 2.0 (1 - 15) | 2.0 (1 - 15) | 4.0 (1 - 13) | **0.0030** |
| Proven (vs. presumed) COVID-19 (n, %) | 77 (97.5) | 51 (96.2) | 26 (100) | 1.000 |
| Haemoglobin (mmol/l)* | 7.4 (6.5 - 8.2)<sup>a</sup> | 7.9 (6.6 - 8.4)<sup>b</sup> | 6.7 (5.9 - 7.3) | **0.0004** |
| WBC (*10<sup>9</sup>/l) | 6.2 (4.3 - 9.7) | 5.8 (3.8 - 9.2) | 8.0 (5.1 - 10.7) | 0.0823 |
| Thrombocytes (*10<sup>9</sup>/l) | 196.5 (136.8 - 278)<sup>c</sup> | 185.5 (122.8 - 278)<sup>b</sup> | 228 (154.3 - 278)<sup>d</sup> | 0.3773 |
| Neutrophils (*10<sup>9</sup>/l) | 4.5 (3.2 - 7.5)<sup>e</sup> | 3.6 (3.0 - 5.3)<sup>f</sup> | 7.3 (4.1 - 9.3) | **0.0024** |
| Lymphocytes (*10<sup>9</sup>/l) | 0.7 (0.4 - 1.1)<sup>e</sup> | 0.7 (0.4 - 1.1)<sup>f</sup> | 0.6 (0.3 - 0.9) | 0.2758 |
| Monocytes (*10<sup>9</sup>/l) | 0.4 (0.2 - 0.7)<sup>e</sup> | 0.4 (0.2 - 0.7)<sup>f</sup> | 0.6 (0.2 - 0.8) | 0.3894 |
| Eosinophils (*10<sup>9</sup>/l) | 0.01 (0.0 - 0.07)<sup>e</sup> | 0.01 (0.0 - 0.03)<sup>f</sup> | 0.04 (0.0 - 0.15) | **0.0150** |
| Basophils (*10<sup>9</sup>/l) | 0.01 (0.0 - 0.02)<sup>e</sup> | 0.01 (0.0 - 0.02)<sup>f</sup> | 0.01 (0.0 - 0.03) | 0.1534 |
| Creatinin (µmol/l) | 83.5 (70.3 - 101.8)<sup>c</sup> | 83 (71 - 102)<sup>g</sup> | 85 (70 - 114.5)<sup>h</sup> | 0.8857 |
| ALAT (U/l) | 33 (21 - 52.5)<sup>i</sup> | 31 (21 - 53)<sup>g</sup> | 34 (21 - 49.3) | 0.6316 |
| CRP (mg/l) | 120 (58 - 232) | 79 (43 - 139.5) | 266.5 (149.8 - 308.5) | **< 0.0001** |
| Ferritin (µg/l) | 1117 (628 - 1780)<sup>a</sup> | 991 (566.5 - 1542)<sup>b</sup> | 1470 (747.8 - 1965) | 0.0557 |
| D-dimer (ng/ml) | 1675 (865 - 2498)<sup>j</sup> | 1150 (760 - 1750)<sup>k</sup> | 3420 (1890 - 6805)<sup>h</sup> | **< 0.0001** |

All values are expressed as median with interquartile ranges (IQR), unless otherwise stated;
ICU: Intensive care unit; WBC: White blood cell count; ALAT: Alanine aminotransferase;
CRP: C-reactive protein
<sup>a</sup>: n = 78. <sup>b</sup>n = 52. <sup>c</sup>: n = 76. <sup>d</sup>: n = 24. <sup>e</sup>: n = 69. <sup>f</sup>: n = 43. <sup>g</sup>: n = 51. <sup>h</sup>: n = 25. <sup>i</sup>: n = 77. <sup>j</sup>: n =
72. <sup>k</sup>: n = 47

### Cytokine concentrations and complement activation in COVID-19 infection

Plasma cytokine measurements showed that IL-6 concentrations were elevated, especially in patients admitted to the ICU (ICU vs. non ICU, median 174.5 pg/ml [IQR 94.5-376.3] vs. 40.0 pg/ml [16.5-81.0], *p* < 0.0001 for day 4-6). In contrast, circulating TNF-α concentrations in COVID-19 patients were low and showed no significant difference between ICU and non ICU patients early in disease (median 24.0 pg/ml [IQR 16.5-33.5] vs. 21.5 pg/ml [IQR 16.0-33.5], *p* = 0.5733 for day 4-6; Figure 1A). Sequential sampling showed that TNF-α remained low during admission with few differences between patients in the ICU or on the ward, with the exception of later during infection when ICU patients had higher concentrations. IL-6 concentrations declined over time but remained high after 10 days in patients primarily admitted to the ICU (Figure 1A). Complement activation was investigated in 78 patients by measuring C3a and terminal complement complex (TCC) (see Supplementary Table 1 for patients characteristics). COVID-19 patients displayed increased activation of complement as compared to healthy controls (HC; n = 10): significantly higher C3a concentrations were demonstrated in ICU (median 556.0 ng/ml [IQR 333.3-712.5]) and non ICU patients (266.5 ng/ml [IQR 191.5-384.0]) as compared to HC (66.5 ng/ml [IQR 60.3-76.0], *p* < 0.05 for both comparisons) at the time of first blood collection, as well as higher TCC concentrations in ICU patients (median 4506 mAU/ml [IQR 3661-6595] vs. 2968 mAU/ml [IQR 2677-3434] in HC, *p* < 0.05). TCC concentrations were not significantly different between non ICU patients (median 3582 mAU/ml [IQR 2947-4300]) and HC. Patients in the ICU had significantly higher plasma C3a and TCC concentrations as compared to non ICU patients (*p* < 0.05 for both components). However, complement activation in both patient groups was less strongly increased compared to patients with bacterial sepsis (median values of C3a 7847 ng/ml [IQR 3996-14408] and TCC 6596 mAU/ml [IQR 5372-15286]; Figure 1B).

**Figure 1.**
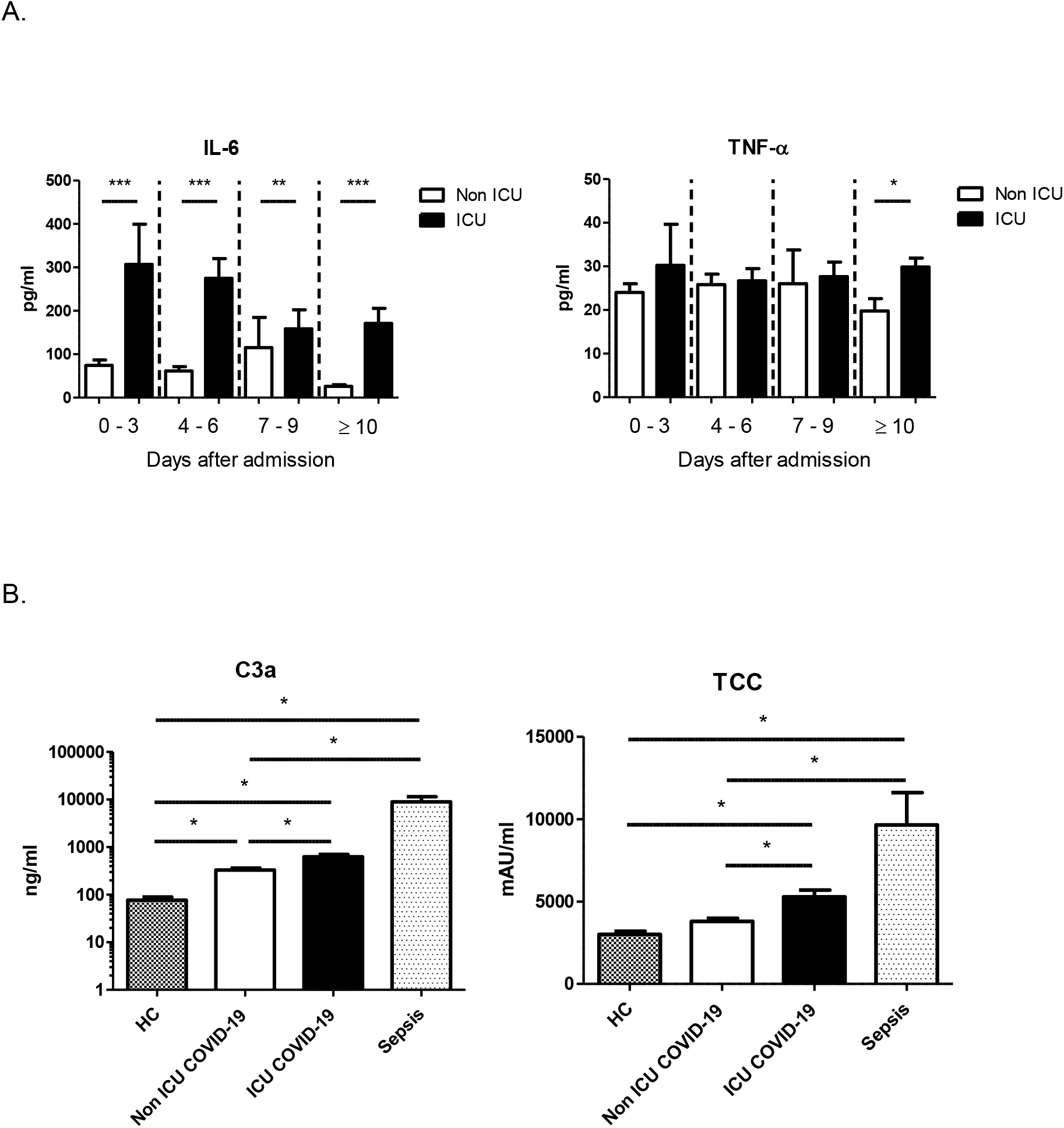
A. TNF-α and IL-6 concentrations in plasma according to time after admission. Comparisons between non-ICU and ICU groups were made by Mann-Whitney test. Bars represent means with SEM. *: *p* < 0.05; **: *p* < 0.01; ***: *p* < 0.0001 TNF-α: Days 0-3: n = 38 (non ICU) and n = 9 (ICU); Days 4-6: n = 22 (non ICU) and n = 17 (ICU); Days 7-9: n = 4 (non ICU) and n = 9 (ICU); > 10 Days: n = 4 (non ICU) and n = 6 (ICU); IL-6: Days 0-3: n = 75 (non ICU) and n = 16 (ICU); Days 4-6: n = 65 (non ICU) and n = 30 (ICU); Days 7-9: n = 21 (non ICU) and n = 23 (ICU); > 10 Days: n = 20 (non ICU) and n = 42 (ICU) B. Terminal complement complex (TCC) and C3a concentrations in plasma at the first time of blood collection. Comparisons between groups were made by Kruskal-Wallis test with Dunn’s multiple comparison test for differences between individual groups. Bars represent means with SEM. For TCC and C3a, p < 0.0001 for the Kruskal-Wallis test. *: *p* < 0.05. HC = healthy controls. n = 10 (HC), n = 52 (non ICU COVID-19), n = 26 (ICU COVID-19), n = 9 (TCC sepsis) and n = 6 (C3a sepsis)

### Inflammatory and cardiometabolic profiling in patients with COVID-19

To perform a comprehensive assessment of inflammatory biomarkers and pathways relevant to COVID-19, we used the proximity extension assay (PEA) based immunoassay (Olink platform) to measure approximately 269 plasma biomarkers in COVID-19 patients (19 ICU versus 28 non ICU patients), sequentially included in our study (see Supplementary Table 2 for patient characteristics). Figure 2 shows that IL-6 (adjusted *p* value 0.001, log fold change (logFC) 2.2) and several chemokines are the most significantly elevated markers in patients with severe COVID-19 in the ICU as compared to non ICU patients. Strikingly, the most downregulated biomarker (with the lowest fold change difference) in patients with severe COVID-19 was stem cell factor (SCF) (adjusted *p* value 0.001, logFC-1.3), a crucial factor for the homeostasis of haematopoiesis.^10^ In contrast, hepatocyte growth factor (HGF) (adjusted *p* value 0.004, logFC 1.4) was significantly higher in ICU patients as compared to non ICU patients. A TNF receptor superfamily ligand (TRAIL) and two receptors (TWEAK, TRANCE) that play a role in apoptosis were significantly lower in patients with severe disease (adjusted *p* value 0.01). Cardiometabolic profiling demonstrated significantly lower dipeptidyl peptidase 4 (DPP4) (adjusted *p* value 0.02, logFC −0.4) and protein C inhibitor (PCI, Serpina5) (adjusted *p* value 0.007, logFC −1.0). They both have a function in regulating the kinin-kallikrein system, in which DPP4 degradates bradykinin and Serpina5 inhibits plasma kallikrein,^11,12^ the enzyme that processes kininogen into bradykinin.

**Figure 2.**
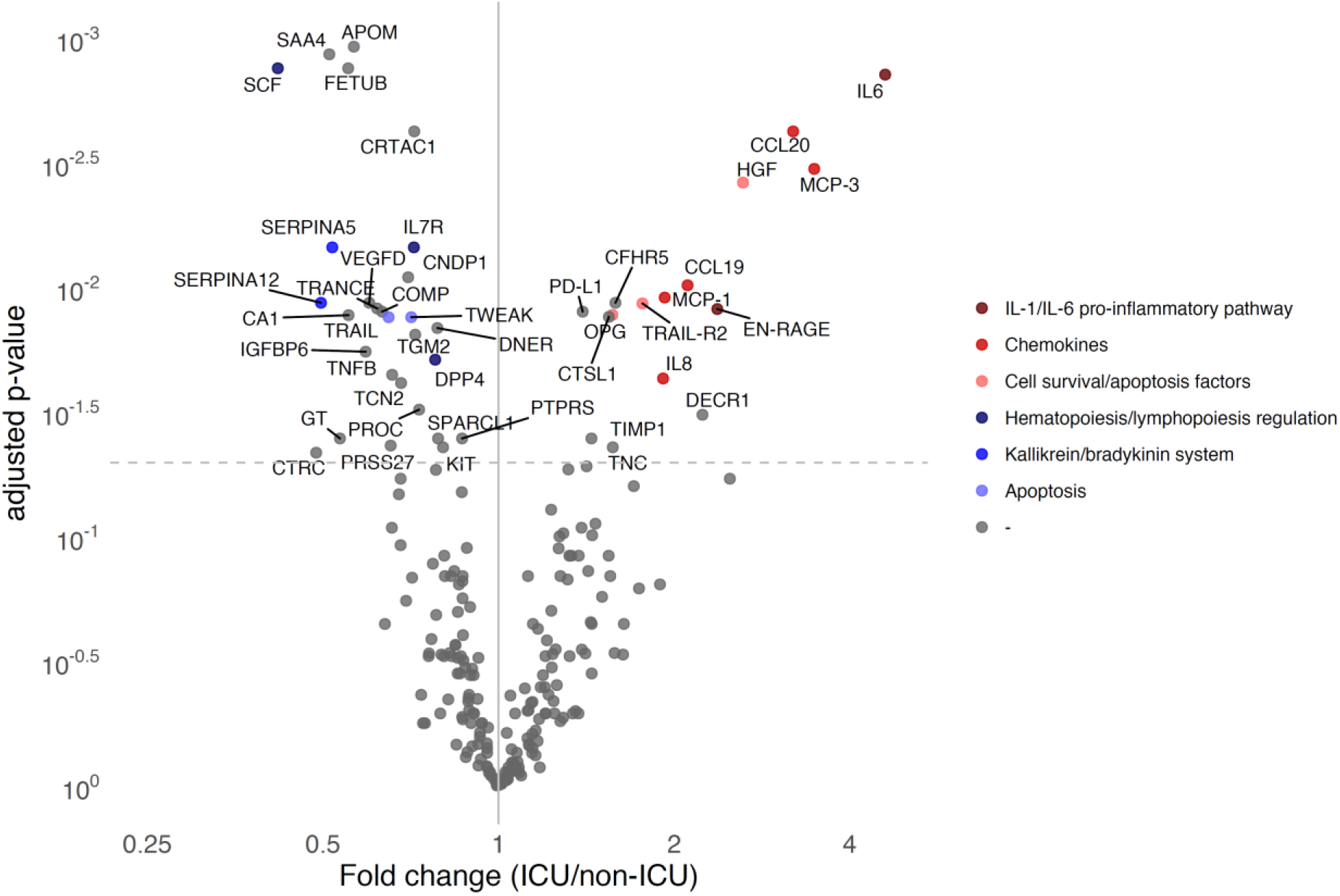
Volcano plot of circulating proteins (n = 235) showing significantly differentially expressed proteins between ICU (n = 19) and non ICU patients (n = 28). Benjamini-Hochberg method used to correct for multiple testing, and adjusted *p* values < 0.05 were considered significant. Age and sex are used as covariates.

### Inflammatory endotypes in COVID-19 patients

Patients with severe infectious diseases such as sepsis can be categorised into immune endotypes that differ in characteristics, trajectories and outcome.^7^ This is important because these endotypes indicate involvement of different pathophysiological mechanisms, which may require different immunomodulatory treatment strategies. Unsupervised clustering analysis of the PEA proteins that significantly differ between ICU and non ICU, C-reactive protein (CRP), D-dimer, ferritin, C3a, C3c and TCC, revealed that ICU patients cluster separately from non ICU patients, but that within these clusters no significantly different profiles could be identified (Figure 3A). All COVID-19 patients have the same profile of markers, which is more pronounced in ICU patients. This indicates that COVID-19 is characterised by a homogeneous inflammatory response and that specific endotypes cannot be discerned. Patients cluster according to disease severity but they all seem to share the same underlying pathophysiological mechanism: activated complement system, an imbalanced kinin-kallikrein system, increased inflammation, lymphopenia, and decreased apoptosis. Although we did not demonstrate any endotypes related to disease severity, there are clear risk factors for severity of COVID-19. We compared men and women admitted to non ICU wards (Figure 3B). Among the differentially expressed inflammatory biomarkers, Serpina12, which is also called vaspin and is able to inhibit tissue kallikreins was lower in men compared to women.^13^ Serum amyloid A4, an acute phase protein with known roles in autoinflammatory syndromes, was also strongly decreased in men compared to women. Interestingly, circulating angiotensin converting enzyme (ACE) 2, which is also the SARS-CoV-2 receptor, was higher in men.

**Figure 3.**
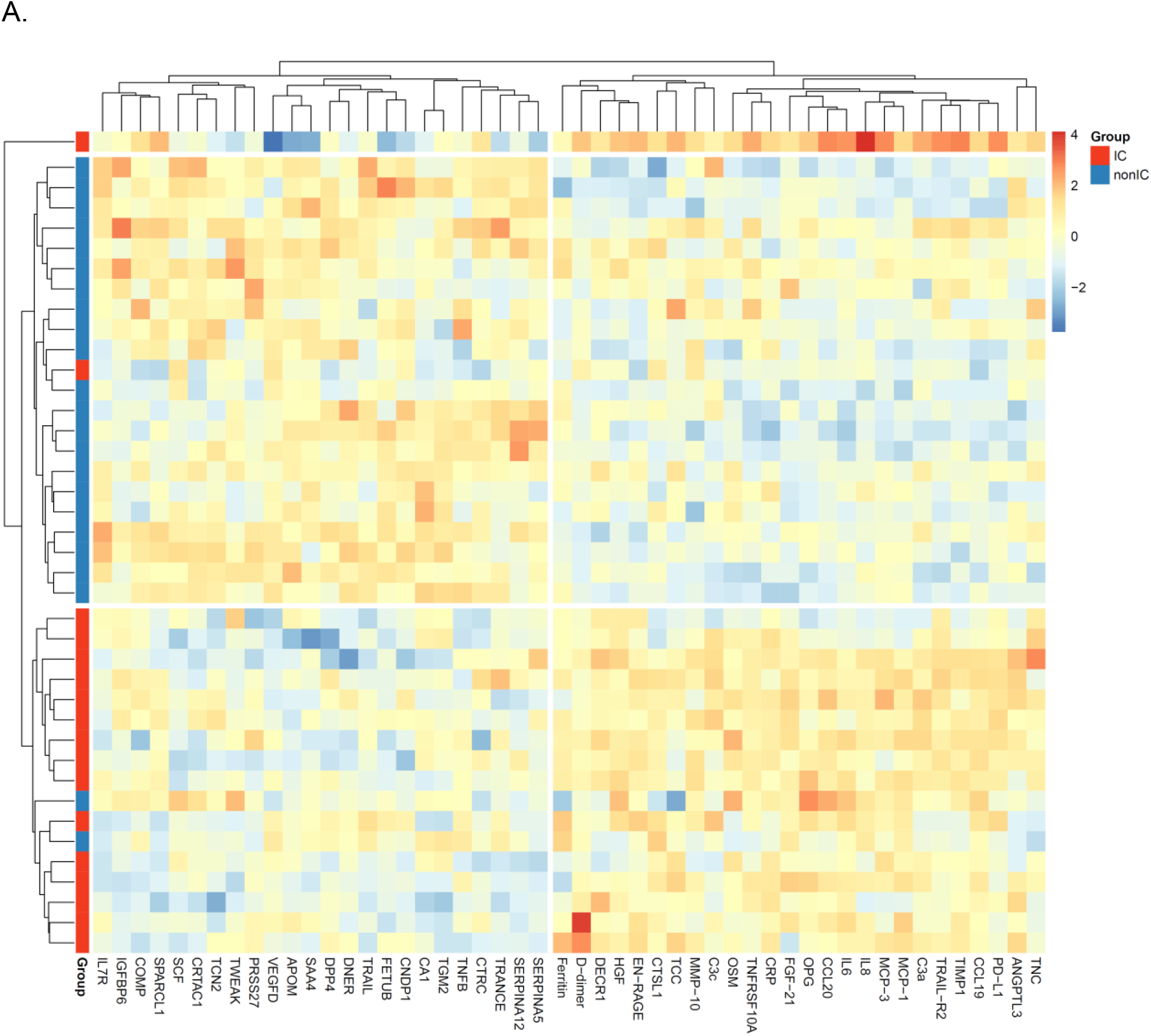
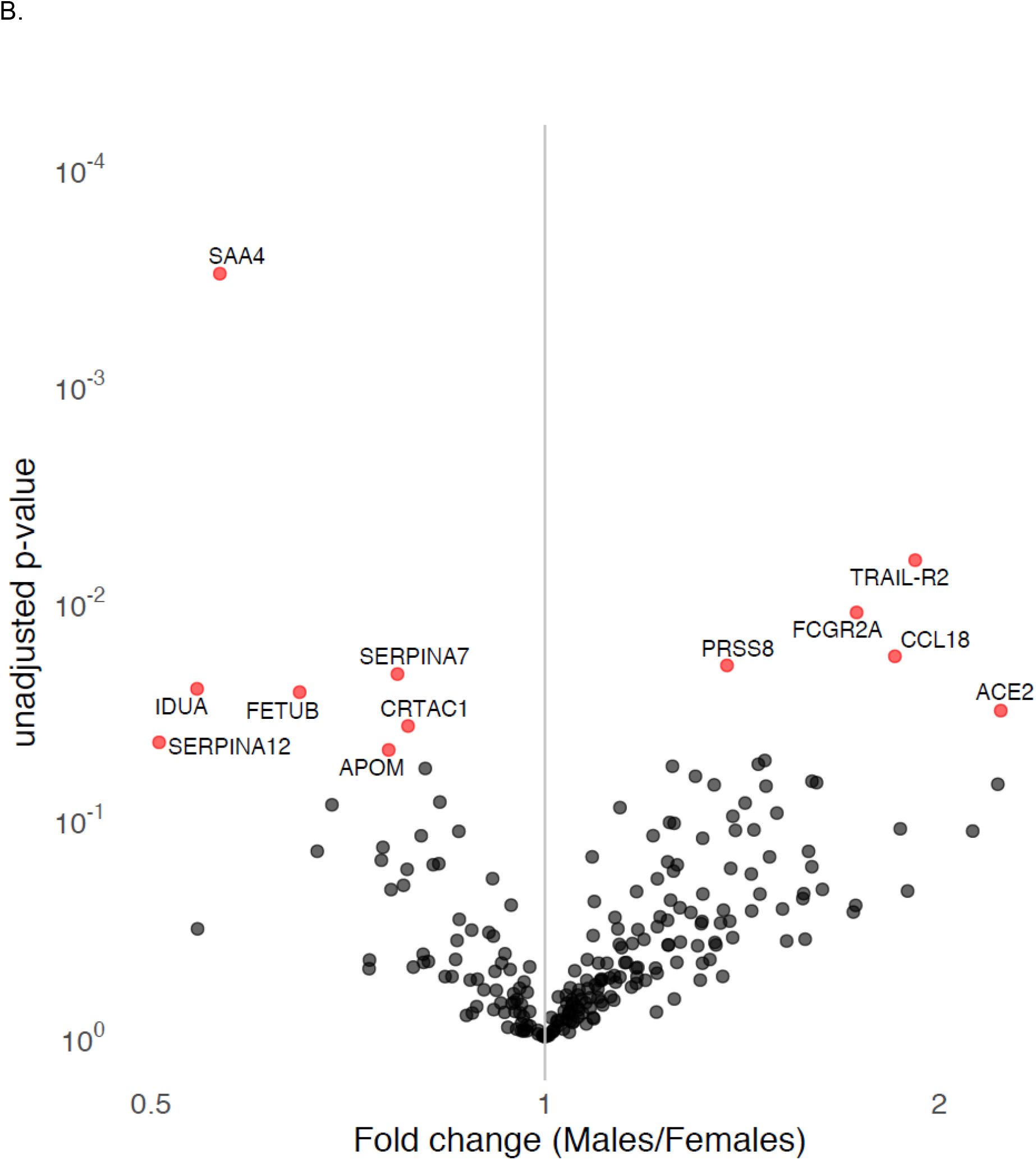
A. Unsupervised hierarchical clustering of protein measurements in ICU patients (n = 17) versus non ICU patients (n = 23) revealed distinct clustering patterns based on disease severity. B. Volcano plot of circulatory proteins (n = 234) of COVID-19 patients on the non ICU ward compared between males (n = 16) and females (n = 12). Differential expression was performed using a linear model with age as covariate, *p* values < 0.05 were considered statistically significant (depicted in red).

## Discussion

Although hyperinflammation is a constant feature of severe infections and sepsis, some clinical characteristics of COVID-19 made us hypothesise that the inflammatory reaction during infection with SARS-CoV-2 also has important particularities that distinguish it from these disease entities: the absence of major haemodynamic consequences such as hypotension, the localised lung edema with the absence of systemic leakage, and the peculiar inflammatory pattern for a viral infection with very high CRP, D-dimers and lymphopenia. We thus hypothesised that the inflammatory reaction in COVID-19 is different from other severe infections.

The assessment of the systemic inflammation in COVID-19 showed that inflammatory markers such as proinflammatory cytokines and complement factors are increased in severely ill COVID-19 patients compared with patients admitted to non ICU wards. The strong increase in IL-6 production, the very high CRP concentrations, and the presence of immature neutrophils in the blood differentiation, all suggest a significant activation of the IL-1 pathway. In contrast, TNF-α circulating concentrations were not strongly induced: this may explain the absence of major systemic vascular dysfunction, for which both IL-1 and TNF-α acting in synergism are needed.^14^ Additional analysis of more biomarkers by Olink technology revealed a number of important pathways that are strongly affected in the severely ill patients: proinflammatory cytokines from the IL-1/IL-6 pathway, anti-apoptotic and proliferative factors, complement, and the kinin-kallikrein system. These data provide strong support for the current clinical trials with both the anti-IL-6 receptor monoclonal antibody tocilizumab and the recombinant human IL-1 receptor antagonist anakinra, of which the results are eagerly awaited.

IL-6 is also an inducer of hepatocyte growth factor (HGF),^15,16^ another cytokine strongly upregulated in critically ill COVID-19 patients. HGF is secreted by mesenchymal cells and acts as a multi-functional cytokine on cells of mainly epithelial origin, in which it regulates cell growth, morphogenesis and tissue regeneration.^17^ Interestingly, recent studies have shown that HGF induces cMET through its receptor, a pathway that is important for plasma cell generation in multiple myeloma.^18^ This observation is paralleled by findings of large numbers of plasma cells in the circulation of COVID-19 patients, as well as in the lungs, where they induce plasma cell endothelitis (Kathrien Grunberg, personal communcation). HGF’s anti-apoptotic and proliferative effects may also play a role in the long-term fibrotic complications in some patients. Other pro-survival metabolic mediators such as FGF21 may also play a role in these processes.

One of the most exciting findings of our analyses is that of the factors involved in the kinin-kallikrein system, which plays an important role in the local inflammation in the lung.^19^ ACE/ACE2 and DPP4 are important enzymes in the degradation pathway of bradykinin, a nonapeptide that regulates vascular permeability. We have recently hypothesised that the loss of bradykinin degradation capacity is a crucial mechanism leading to pulmonary angioedema in COVID-19.^20^ Moreover, we now demonstrate that Serpina5, an inhibitor of plasma kallikrein and DPP4, which degradates bradykinin, are significantly lower in severe COVID-19 disease. Plasma kallikrein processes high molecular weight kininogen (HMWK) into bradykinin, which in turn will activate bradykinin receptor 2 (B2R) that is constitutively expressed on endothelial cells in the lung. In addition, tissue kallikrein can also contribute to local bradykinin formation, and we observed that Serpina12, which is a specific tissue kallikrein inhibitor, was lower in men. The vicious cycle of an activated kinin-kallikrein system resulting in bradykinin receptor activation due to loss of inhibitory enzymes is key for the vascular leakage. The kinin-kallikrein system may thus represent an important therapeutic target in severe COVID-19 with ARDS, and proof-of-principle clinical trials are currently under way to test this hypothesis in our institution.

In addition to the inflammatory factors that are upregulated in COVID-19 patients in the ICU, a number of cytokines were shown to be lower in the severely ill patients. Among them, most notable is the strong decrease in SCF. SCF (also known as KIT-ligand) is a cytokine that binds to the c-KIT receptor (CD117), and plays an important role in the regulation of haematopoietic stem cells (HSCs) in the stem cell niche in the bone marrow.^10^ SCF stimulates the survival of HSCs in vitro and induces self-brenewal and maintenance of HSCs *in vivo.^21^* It is thus tempting to speculate that the strong downregulation of SCF in patients with severe forms of COVID-19 contributes to the deep and sustained lymphopenia that accompanies a poor outcome.^22^

Adjuvant host-directed therapies in severe infections such as sepsis have been proposed to have the potential to improve the outcome of patients. However, all immunotherapies investigated in sepsis in the last three decades failed to show clinical efficacy, and it has been hypothesised that the lack of adjustment of the immunotherapy approach to the (specific) immune status of the patient is one of the most important reasons for this.^23^ Sepsis endotypes based on transcriptional patterns in circulating immune cells have been described to influence patient outcomes,^7^ and clinical trials have been designed to treat patients in a personalised approach. We also investigated whether we could identify inflammatory endotypes among COVID-19 patients based on the comprehensive assessment of inflammatory markers measured: one could envisage that the pathophysiology of the disease in some patients would be characterised by excessive activation of the IL-1/IL-6 pathway, while in other patients disease would be mainly caused by the kinin-kallikrein system or complement activation. However, unbiased clustering of COVID-19 patients differentiated patients based on disease severity (ICU versus non ICU), rather than identifying different inflammatory clusters (Figure 2). This suggest a relative homogeneity of the inflammatory pathophysiology of the patients. We cannot exclude late differentiation of patients more prone to specific complications (e.g., late progression to fibrosis), but these current insights suggest that the inflammation in the majority of patients follow a relatively homogeneous pattern which can be used as a guide for therapy.

All these data allow to build a pathogenetic model of inflammation in COVID-19 patients, which might guide immunotherapeutic approaches with the highest potential to translate into clinical benefit. In the beginning of the SARS-CoV-2 infection, a broad activation of innate immunity mechanisms is induced by the virus, which is necessary for the induction of host defense and virus elimination. While this is successful in the majority of patients, in a significant minority of them the disease progresses to a more severe form necessitating ICU admission.

In conclusion, the present study is the first comprehensive assessment of inflammatory pathways in COVID-19 patients (Figure 4). The main pathways of dysregulation of inflammation are described that correlate with increased severity, including an unknown role for the kinin-kallikrein system and depression of stem cell factor as a likely contributor to lymphopenia. Future studies are needed to engage these pathways therapeutically, and to attempt to improve the outcome of severely ill patients with COVID-19.

**Figure 4.**
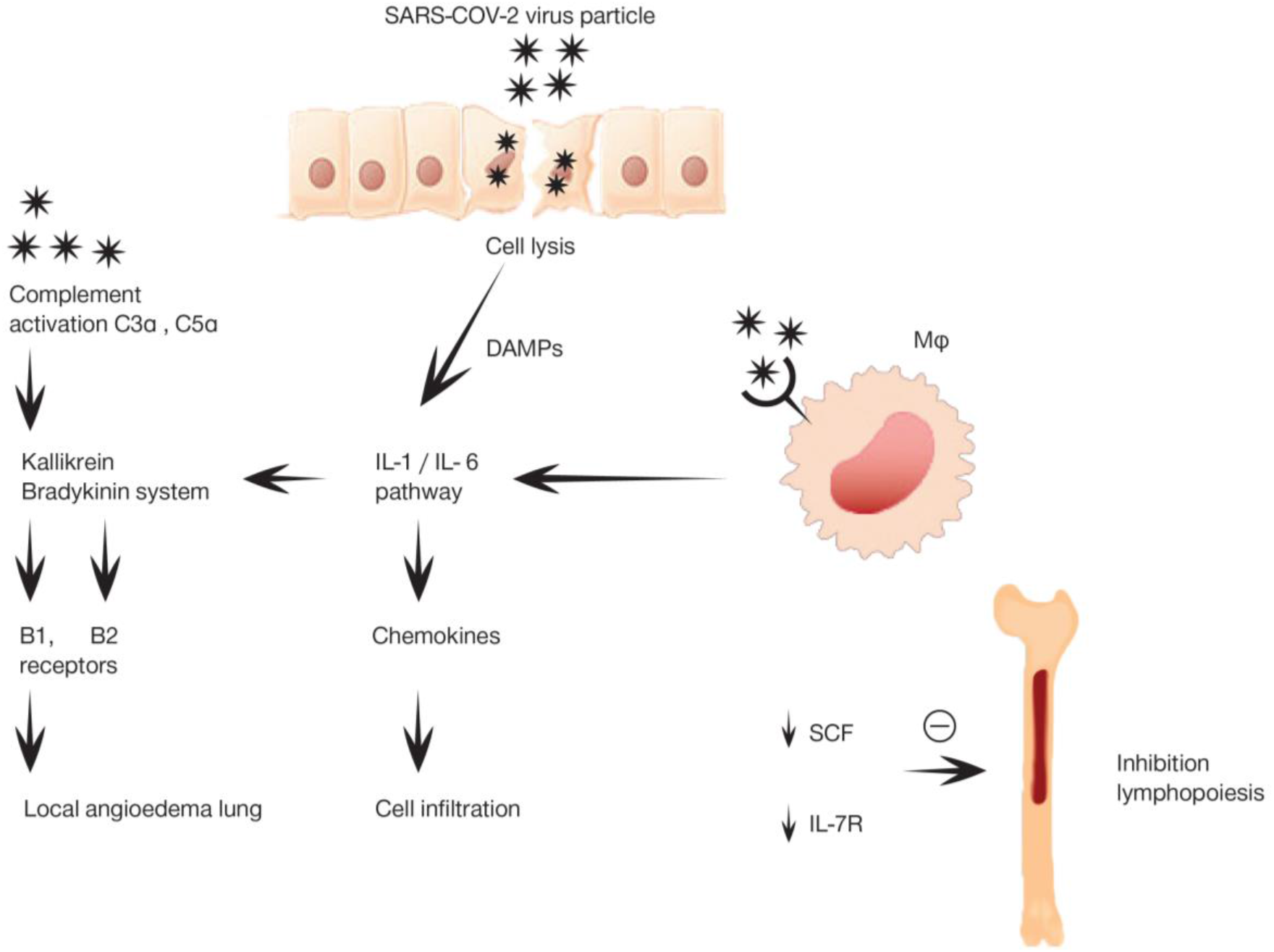

## Data Availability

All data are available for review upon request.

## Acknowledgments

The authors would like to thank the entire RCI-COVID-19 study group: Martin Jaeger, Helga Dijkstra, Heidi Lemmers, Liesbeth van Emst, Kiki Schraa, Cor Jacobs, Anneke Hijmans, Trees Jansen, Fieke Weren, Liz Fransman, Jelle Gerretsen, Hetty van der Eng, Noortje Rovers, Margreet Klop-Riehl, Josephine van de Maat, Gerine Nijman, Simone Moorlag, Esther Taks, Priya Debisarun, Heiman Wertheim, Joost Hopman, Janette Rahamat-Langendoen, Chantal Bleeker-Rovers, Jaap ten Oever, Esther Fasse, Esther van Rijssen, Manon Kolkman, Bram van Cranenbroek, Pleun Hemelaar, Remi Beunders, Sjef van der Velde, Emma Kooistra, Nicole Waalders, Wout Claassen, Hidde Heesakkers, Tirsa van Schaik. All of these authors are affiliated to the Radboud Center for Infectious Diseases.

The authors want to thank Olink Proteomics AB (Uppsala Sweden) for their donation of multiplex proximity extension assays.

FLvdV was supported by a Vidi grant of the Netherlands Association for Scientific Research. MGN was supported by an ERC Advanced grant (#833247) and a Spinoza Grant of the Netherlands Association for Scientific Research.

## Author contributions

Conceptualisation, FLvdV., MvD, JWMvdM, LABJ. and MGN; Formal analysis, NAFJ, IG, AHdN, VACMK, VM, CKB and VK; Investigation, FLvdV, NAFJ, IG, AHdN, ET, AHS, IEH, LABJ and MGN; Resources, FLvdV, NAFJ, IG, AHdN, MK, HJPMK, RLS, IJEK, HvdH, JAS, TF, MR, WH, TTSMD, APMK, KAS, KV, CM, AHS, IEH, LW, ET, EJG-B, LABJ, MMvdH, JH, QdM, PP and MGN; Data curation, NAFJ, IG and AHdN; Writing – Original draft: FLvdV and MGN; Writing – Review & editing: FLvdV, NAFJ, IG, AHdN, VACMK, VM, HJPMK, RLS, IJ, RJMB, IJEK, HvdH, JAS, TF, MR, WH, TTSMD, APMK, KAS, KV, CM, AHS, IEH, LPGD, LW, ET, MvD, JWMvdM, RvC, EJG-B, LABJ, MMvdH, JH, QdM, PP and MGN; Supervision, FLvdV, LABJ and MGN; Project administration, FLvdV, NAFJ, IG, AHdN, LABJ and MGN; Funding acquisition: FLvdV, LABJ and MGN.

